# Health economic simulation modeling of an AI-enabled clinical decision support system for coronary revascularization

**DOI:** 10.1101/2025.05.28.25328522

**Authors:** Tom Mullie, Arjun Puri, Emma Bogner, Bryan Har, Colm J. Murphy, Robert C. Welsh, Benjamin Tyrrell, Christopher L. F. Sun, Joon Lee

**Affiliations:** Acute Care Alberta, Calgary, Alberta, Canada; Data Intelligence for Health Lab, Cumming School of Medicine, University of Calgary, Calgary, Alberta, Canada; Department of Cardiac Sciences, Cumming School of Medicine, University of Calgary, Calgary, Alberta, Canada; Libin Cardiovascular Institute, Cumming School of Medicine, University of Calgary, Calgary, Alberta, Canada; Golden State Heart and Vascular, Burlingame, California, United States; Mazankowski Alberta Heart Institute and University of Alberta, Division of Cardiology, Edmonton, Alberta, Canada; Royal Alexandra Hospital, Edmonton, Alberta, Canada; Telfer School of Management, University of Ottawa, Ottawa, Ontario, Canada; University of Ottawa Heart Institute, Ottawa, Ontario, Canada; Department of Community Health Sciences, Cumming School of Medicine, University of Calgary, Calgary, Alberta, Canada

## Abstract

**Importance:** Coronary revascularization decision-making can be challenging. While artificial intelligence (AI) models have been developed to support this decision-making, health economic evaluation of such models has been rare.

**Objective:** To evaluate the economic value of an AI-enabled coronary revascularization decision support system in terms of cost savings and gains in quality adjusted life years (QALY).

**Design:** Retrospective health economic simulation modeling study using real-world patient data and AI-generated patient outcome predictions.

**Setting:** 26,605 adult patients with obstructive coronary artery disease who underwent diagnostic coronary angiography between 2009 and 2019 in Alberta, Canada.

**Exposures:** Clinicians deciding among medical therapy only, percutaneous coronary intervention, and coronary artery bypass grafting were simulated to be provided with AI-generated decision support information in the form of 3- and 5-year major adverse cardiovascular event and all-cause mortality predictions.

**Main Outcomes and Measures:** Average cost savings and gains in QALY, represented as a willingness-to-pay, per patient resulting from treatment decisions altered by the AI-generated decision support.

**Results:** Most actual coronary revascularization decisions could have been improved by AI decision support from a health economic perspective. At a willingness-to-pay of $50,000 per QALY, as many as 51% of all actual treatment decisions shifted to another treatment, resulting in an average cost saving of $31,204 and a QALY gain equivalent to up to $2,406 per patient. Even in a conservative scenario where clinicians’ AI adoption was limited by ignoring AI recommendations unless the gain in QALY was substantial, 22.4% of the actual decisions shifted, resulting in an average gain of 0.327 QALY, equivalent to up to $16,371, per patient.

**Conclusions and Relevance:** AI can help clinicians to optimize coronary revascularization decisions. The health system level economic value of optimized treatment decisions can be substantial in the form of reduced costs stemming from fewer future complications and improved patient outcomes.

**Key Points:** *Question:* How much cost saving and gain in quality adjusted life years (QALY) can be expected from using an AI-enabled clinical decision support system for coronary revascularization decision-making?

*Findings:* AI was able to improve the cost-effectiveness of 51% percent of actual treatment decisions. Pursuing AI-based optimal treatments would have resulted in an average cost saving of $31,204 and a QALY gain equivalent to $2,406 per patient.

*Meaning:* AI can help optimize coronary revascularization decisions, leading to substantial economic value in the form of cost savings and improved patient outcomes.

## Introduction

Coronary revascularization decision-making for patients with coronary artery disease (CAD) is often challenging when complex disease and unique patient characteristics preclude a straightforward application of clinical practice guidelines. Previous studies have investigated and discussed the treatment decision-making challenges associated with left-main or multivessel disease, co-morbidities, complex coronary anatomies, and patient preference.^1–4^

When evidence-based coronary revascularization decision-making is difficult, artificial intelligence (AI) can potentially present a viable alternative by providing personalized, data-driven insights. Several studies have demonstrated promising performances of AI models that predicted mortality and major adverse cardiovascular event (MACE) outcomes at various time points.^5–8^

While AI prediction performance has been investigated in the context of CAD and coronary revascularization, little is known about the economic value that AI can potentially bring to CAD care. Given that economic benefits are crucial to AI adoption in healthcare settings, this study aimed to perform health economic analysis of an existing AI-enabled clinical decision support system (CDSS) for coronary revascularization. Using a Markov chain simulation model and retrospective patient data, this study assessed the potential value of improving coronary revascularization decisions among percutaneous coronary intervention (PCI), coronary artery bypass grafting (CABG), and medical therapy only (MT) made by interventional cardiologists following diagnostic coronary angiography.

## Methods

### AI-enabled CDSS

We conducted health economic analysis on an existing AI-enabled CDSS named Revaz AI, the development and prediction performance evaluation of which have been described previously.^9^ In this study, we evaluated the Revaz AI models that predict the likelihoods of 3- and 5-year all-cause mortality and 3-point MACE (heart failure, myocardial infarction [MI], and stroke) post-diagnostic coronary angiography. These models were developed using a comprehensive data set from over 42,000 patients who underwent coronary angiography in Alberta, Canada from 2009 to 2019. They are designed to help decide among PCI, CABG, and MT for patients with obstructive CAD (defined as at least 50% or 70% stenosis in the left main or other coronary vessels, respectively). Patients presenting with ST elevation MI (STEMI) are ineligible for Revaz AI due to its emergent nature that requires immediate PCI.

### Patient Cohort

Our economic analysis was based on real-world data from 26,605 patients who underwent diagnostic coronary angiography in Alberta, Canada between 2009 and 2019. This was a subset of the data set used to develop Revaz AI with the following inclusion criteria: 1) 5-year follow-up to enable all outcome predictions, 2) age ≥ 18 years, and 3) presentation of stable angina, unstable angina, or non-ST elevation MI (NSTEMI). eTable 1 (Supplement 1) tabulates the patient characteristics used in the simulation model.

### Health Economic Simulation Model

The simulation model assumed that patients undergoing diagnostic coronary angiography have the possibility of receiving any of the three treatment options: PCI, CABG, and MT. In practice, as shown in eFigure 1 (Supplement 1), there would be multiple decision points and factors which might influence treatment choice. For simplicity, however, this study condensed the decision process to one hypothetical point in which diagnostic angiography has been performed and a treatment decision must now be made.

For each patient in the data set, Revaz AI predicted a set of outcome probabilities conditioned on treatment. Those outcome probabilities were then fed to a Markov simulation model (see eFigure 2 [Supplement 1] for the model structure) with a quarter year cycle length and a lifetime horizon. Each of the three treatment paths was simulated 500 times with the same base outcome probabilities. Patients entered the model at their actual age at the time of angiography and with their actual indication for angiography (stable angina, unstable angina, or NSTEMI) and extent of CAD (single vessel [SV], multivessel [MV], or left main [LM]). Event probabilities not directly taken from Revaz AI were extracted from the literature and are shown in eTable 2 (Supplement 1).

The full technical details of the simulation model are described in Supplement 2.

### Costs

eTable 3 (Supplement 1) reports all costs used in the simulation model in 2024 US dollars. Where necessary, costs were converted to US dollars at purchasing power parity^10^ and inflated to 2024 prices using the Bureau of Labor Statistics’ Consumer Price Index for Medical Care.^11^

Initial treatment cost and the cost of repeat revascularization during follow-up were based on trial data averaged across the EXCEL, SYNTAX, and FREEDOM trials.^12^ MT patients were assumed to receive diagnostic catheterization. Additionally, PCI procedures incurred a consumable cost for second generation drug eluting stents, with the number of stents determined by the type of occlusion. Acute procedure costs were assumed to be gamma distributed, with variance equal to 5 times the mean. Values were simulated at the individual procedure level.

Ongoing post-procedure management costs were taken from the literature and included outpatient care, rehabilitation where necessary, and physician fees. Additionally, patients were assumed to follow a drug regimen for the remainder of their lives based on their diagnosis and treatment. Patients who presented with stable angina and received revascularization were assigned aspirin 325mg, a statin (atorvastatin 40mg), and an ACE inhibitor (lisinopril 5mg). If those patients were assigned to MT, a beta blocker (metoprolol 100mg) and a calcium channel blocker (amlodipine 5mg) were added. For patients who presented with unstable angina or NSTEMI, regardless of treatment, clopidogrel 75mg was added.

Patients who suffered a MACE had additional costs applied. Each MACE had an initial cost of treatment, as well as an ongoing follow-up cost for the remainder of the patient’s life. Patients could suffer repeated MACEs of the same type and would accrue acute treatment costs each time, but ongoing costs did not increase with repeat episodes of the same MACE type.

All ongoing costs varied randomly at the simulation level. Draws were normally distributed with a standard deviation equal to 10% of the mean. Half-cycle corrections were applied to ongoing costs in cycles where the patient died.

### Utilities

Health related quality of life (HRQoL) was estimated based on a model calibrated using the English general population.^13^ Base HRQoL utility was estimated based on patient age and sex, representing a typical utility value for a healthy adult (eFigure 3 [Supplement 1]). Modifiers were then applied based on the individual’s health conditions and treatment history. Patients could have multiple modifiers applied at once to reflect a history of multiple conditions and the impact of treatment. Cardiac health states were assumed to include MT, and no treatment modifier was applied for patients receiving MT for their condition.

The base utility was multiplied by modifiers indicating whether the patient had received either PCI or CABG in the cycle before or in the past, relative to expected HRQoL under MT (it was assumed that no patient received no intervention). Modifiers were based on reported changes in HRQoL post-treatment for patients in the EXCEL trial.^14^ eTable 4 (Supplement 1) summarizes the utility modifiers used in the model.

Base utilities were retained across simulations, but health state modifiers were randomly simulated with a normal distribution and standard deviation equal to 1% of mean. Treatment modifiers were simulated with a normal distribution and a standard deviation equal to 0.5% of the mean.

### Simulation Scenarios

Three scenarios were simulated, where Scenario 1 was the primary analysis, and Scenarios 2 and 3 were secondary analyses. In Scenario 1, physicians were assumed to make the health economically optimal decision by maximizing patient outcomes while adhering to a maximum willingness-to-pay threshold of $50,000 per quality adjusted life year (QALY), a commonly used threshold in health care cost-effectiveness analysis.^15^

Scenario 2 assumed that physicians maximized patient expected QALY with no cost threshold, because, in practice, physicians are likely less sensitive to health economic measures and primarily interested in maximizing benefit for their individual patients.

Scenario 3 was an investigation of AI adoption where physicians pursued their originally selected treatment unless Revaz AI recommended an alternative which added at least 0.2 QALY to the patient’s expected rest of life HRQoL. As in Scenario 2, physicians never opted for a treatment that resulted in worse outcomes.

### Research Ethics and Reporting Guidelines

This study was approved by the Conjoint Health Research Ethics Board at the University of Calgary (REB20-1879). This study followed the Consolidated Health Economic Evaluation Reporting Standards (CHEERS) 2022 Statement (see Supplement 3).

## Results

### Primary Analysis - Scenario 1

Table 1 shows that the net impact of pursuing optimal treatment as recommended by Revaz AI would lead to a net increase of 1,716 CABG procedures, with a decrease in MT (−221) and PCI (−1,495). Overall, 13,055 of 26,605 patients (49%) remained with their initial treatment, while the remainder shifted. It is notable that shifts occurred in all possible directions, with more than 15% of the patients initially assigned to any one treatment being recommended to shift to each alternative.

**Table 1:** Actual treatment compared to economically optimal treatment in Scenario 1. MT: medical therapy only, PCI: percutaneous coronary intervention, CABG: coronary artery bypass graft.

|  |  | **Actual Treatment** |  |  |
| --- | --- | --- | --- | --- |
|  |  | **MT** | **PCI** | **CABG** |
| **Optimal Treatment** | **MT** | 1,733 | 3,315 | 610 |
|  | **PCI** | 3,253 | 10,026 | 2,023 |
|  | **CABG** | 893 | 3,456 | 1,296 |
| **Net Gain/Loss** |  | -221 | -1,495 | +1,716 |

Figure 1 illustrates the net change in system cost and QALY experienced (valued at the maximum willingness-to-pay of $50,000/QALY). The average impact was a cost saving of $31,204 and a QALY gain of 0.048 corresponding to a maximum willingness-to-pay of $2,406. However, the range of shifts was broad and for most patients whose treatment changed the impact was either an increase in costs that was cost effective given an improvement in outcomes or a decrease in costs greater than the willingness-to-pay to avoid a decrease in HRQoL. Overall, applying Revaz AI was dominant over standard care as long as it was cost saving and outcome improving (i.e., at any price below $31,204 in this Scenario since Revaz AI improved outcomes on average).

**Figure 1:**
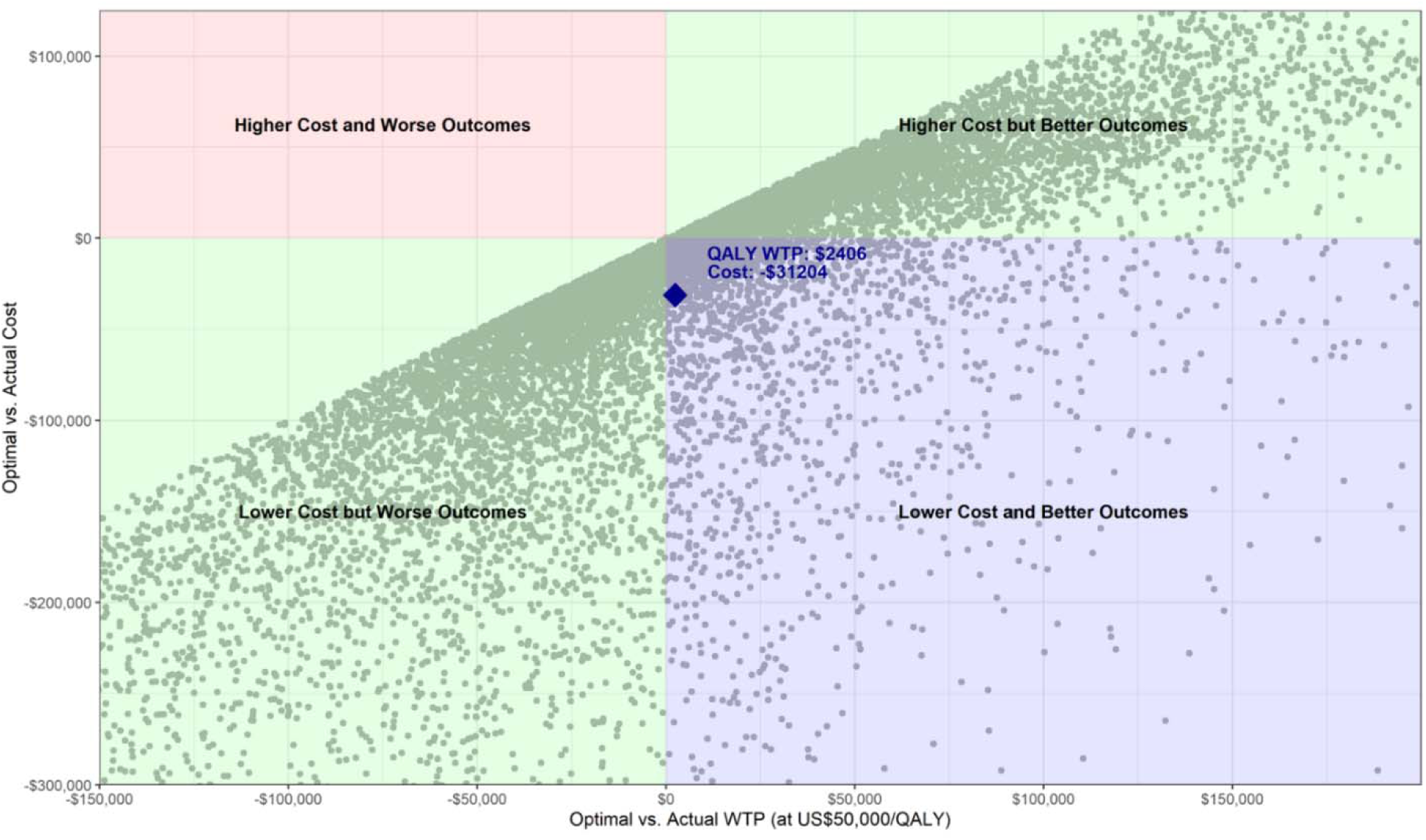
Impact of Revaz AI on individual cost and health related quality of life outcomes in Scenario 1 with a maximum WTP of $50,000/QALY. Each point represents an individual patient. WTP: willingness to pay, QALY: quality adjusted life year.

### Secondary Analyses - Scenarios 2 and 3

Table 2 and Figure 2 present the results from Scenario 2. As with Scenario 1, patients shifted in all possible directions, but the net flow from MT and PCI to CABG was exaggerated. Consequently, costs increased significantly, by $40,163. This included both immediate costs from shifting towards more intensive initial treatment as well as long-term costs due to increased survival. The average QALY gained increased significantly, to 0.61, although on average the shift was not cost effective by conventional thresholds (an incremental cost-effectiveness ratio [ICER] of $65,841/QALY).

**Figure 2:**
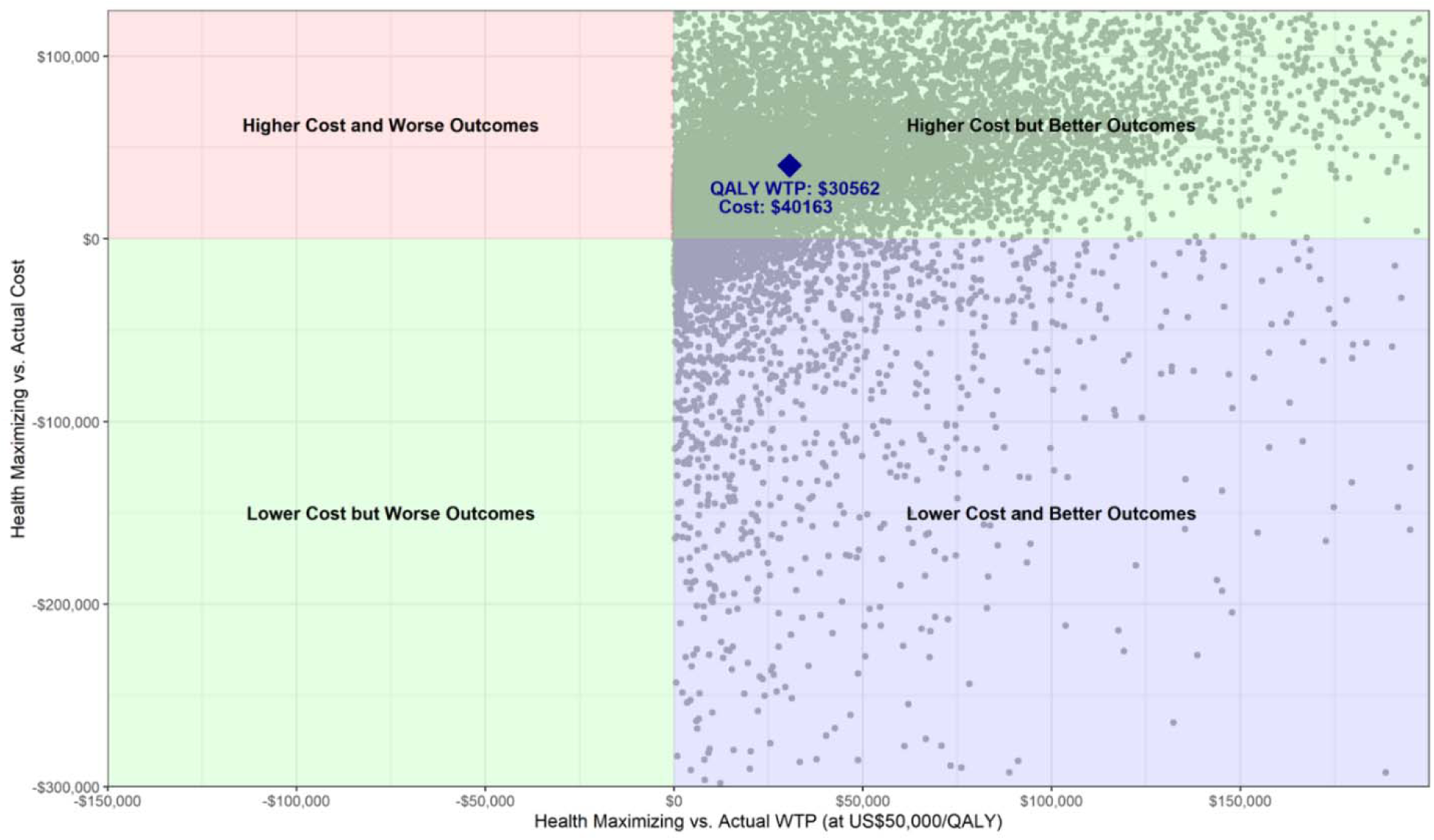
Impact of Revaz AI on individual cost and health related quality of life outcomes in Scenario 2 that maximized QALY without a cost threshold. Each point represents an individual patient. WTP: willingness to pay, QALY: quality adjusted life year.

**Table 2:** Actual treatment compared to QALY maximizing treatment in Scenario 2. MT: medical therapy only, PCI: percutaneous coronary intervention, CABG: coronary artery bypass graft.

|  |  | **Actual Treatment** |  |  |
| --- | --- | --- | --- | --- |
|  |  | **MT** | **PCI** | **CABG** |
| **Optimal Treatment** | **MT** | 432 | 926 | 193 |
|  | **PCI** | 2,335 | 7,973 | 1,452 |
|  | **CABG** | 3,112 | 7,898 | 2,284 |
| **Net Gain/Loss** |  | -4,328 | -5,037 | +9,365 |

eTable 5 (Supplement 1) and Figure 3 show the results from Scenario 3. Most patients retained their original treatment, although as in the other Scenarios there was at least some flow from each alternative treatment path to each of the others. It is notable that even in this conservative Scenario, Revaz AI shifted 5,960 (22.4%) of all treatment decisions.

**Figure 3:**
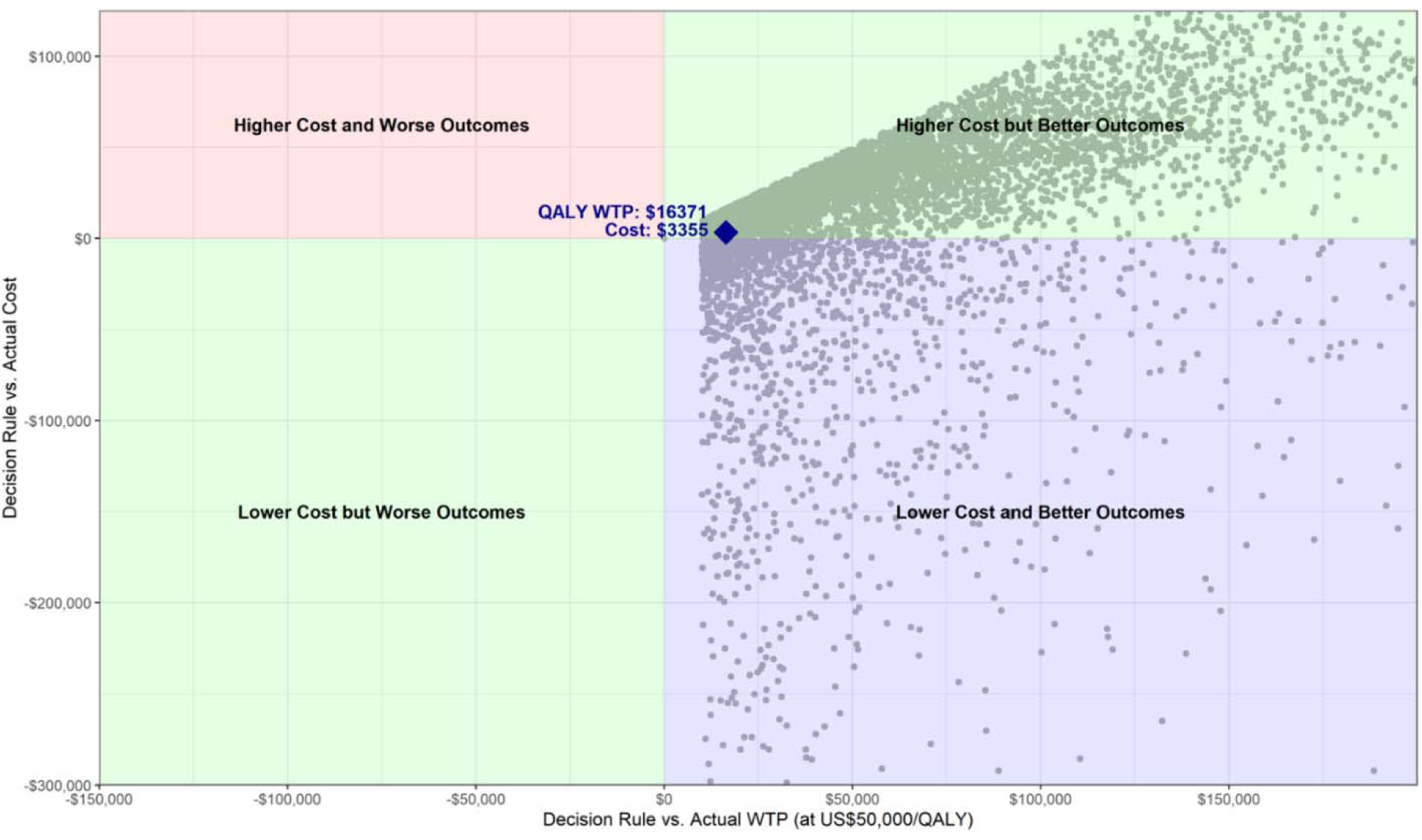
Impact of Revaz AI on individual cost and health related quality of life outcomes in Scenario 3 where physicians followed Revaz AI only when there was a QALY gain of at least 0.2. Each point represents an individual patient. WTP: willingness to pay, QALY: quality adjusted life year.

The decision rule in this Scenario was also cost increasing, although by a much smaller amount ($3,355/patient), and resulted in significant QALY gains (0.327 QALY/patient) compared to strict adherence to the economically optimal treatment (Scenario 1) but less than the impact of simply maximizing QALYs gained (Scenario 2). Applying Revaz AI was cost effective in this scenario, with an ICER of $10,260/QALY.

## Discussion

Our results show that Revaz AI can improve the cost-effectiveness of actual CAD treatments in most cases, and that even with conservative assumptions regarding the probability of switching treatment decisions, more than a fifth of patients would have their course of treatment altered if Revaz AI were applied. The specifics of the shift will depend on the AI adoption criteria by which physicians weigh cost versus HRQoL for their patients, and the extent to which they are, in practice, willing to allow Revaz AI’s recommendations to alter their judgement. Prospective clinical trials are required to determine that willingness in real-world conditions.

Previous Markov simulation-based health economic studies tended to focus on screening tools with a binary diagnostic decision.^16–18^ On the contrary, the present study assessed which of the three competing treatment paths was optimal in the presence of considerable uncertainty. As a result, we opted for extensive empirical simulations for each patient to accurately quantify the economic value of Revaz AI, at the expense of substantially increased computational costs. Our work demonstrates that more sophisticated computational techniques can enable health economic analysis of complex health care technologies such as AI-based clinical decision support.

The economic simulation model used treatment costs faced by a typical US healthcare system, and a standard US willingness to pay threshold of $50,000/QALY was used to represent the cost-HRQoL tradeoff decision-making in the US. Different jurisdictions would have different costs as well as different cost effectiveness thresholds, which would change the optimal therapy as recommended by the model in some cases and would alter the willingness to pay for benefits created. While the $50,000/QALY threshold has been common in the literature, higher thresholds ranging from $100,000 to $150,000/QALY have been increasing used in recent years.^19^

Despite the rapidly increasing interest in AI innovations in healthcare, there has been a paucity of economic evaluations of health AI technologies in the literature.^20,21^ The few studies that have examined the economic value of AI in healthcare tended to focus on medical imaging rather than clinical decision support, and on cost reduction rather than long-term impact.^20^ It has also been acknowledged that health economic evaluation of AI is challenging due to its rapid technological advances, lack of generalizability, need for reconfigured clinical workflows, and potential to exacerbate health disparities.^22^ The present study makes meaningful contributions to this understudied area of health AI research.

### Limitations

This study has several limitations. First and foremost, Revaz AI outcome probabilities were assumed to be a correct representation of the future outcomes for the patient for whom they were generated. Although Revaz AI was trained using a large-scale multi-center data set and its prediction performance is state-of-the-art,^9^ any inaccuracy or bias in Revaz AI predictions would reduce the value of Revaz AI in directing care. Additionally, Revaz AI was trained on a data set where clinicians incorporated comprehensive clinical factors and patient preference to arrive at a defined revascularization strategy, which could influence its applicability in a prospective cohort. Given the reliance on the model’s accuracy, the results presented here should be interpreted as a ceiling for Revaz AI’s potential value.

Second, the results assume that Revaz AI could and would be applied in all cases, and that its optimal treatment recommendations would be followed. In practice, physicians and patients may diverge from Revaz AI’s recommendations for several reasons. For example, from a general health system perspective, Revaz AI recommended an overall shift towards increasing CABG volumes, primarily at the expense of PCI. In practice, this may not be feasible because: 1) the patient may not be a good surgical candidate (e.g., older frail patients) or prefer less invasive procedures; 2) CABG may not be technically feasible due to the coronary anatomy or lesion locations; 3) patients and physicians may choose to weigh the risks of CABG differently than Revaz AI for reasons not discernable in the data ingested by Revaz AI; or 4) the surgical capacity of the health system may be limited. As with the previous limitation, the results from this study should be considered a ceiling, with each case in which an ‘optimal’ strategy is not pursued reducing the potential overall value of Revaz AI.

Third, while the econometric simulation model relied on trial data (primarily SYNTAX,^23^ FREEDOM,^24^ and EXCEL^12^) which covered a broad range of populations, the data on which Revaz AI was trained and the population for whom the benefit estimates were calculated represented specifically the population of the province of Alberta, Canada.

Fourth, the trials on which the simulation model was based are more than a decade old, and the more recent ISCHEMIA trial^25^ has not yet published economic evaluation results. Any significant developments in MT, PCI, or CABG procedures would also change the economic results.

Fifth, as Supplement 2 describes, there were many underlying assumptions in the simulation model. The computational costs associated with the extensive simulation modeling in this study prohibited a complete sensitivity analysis on all assumptions, as each new combination of assumptions and model parameters would require a repeat of the entire simulation.

Lastly, this study only focused on the economic value at the patient level stemming from optimized revascularization decisions. Initial infrastructure investments and operational costs related to using Revaz AI at the point of care could be substantial but were excluded.

All these limitations should be addressed with further research, particularly through clinical trials of Revaz AI to observe real-world impacts on clinical decision-making and any changes in patient outcomes as Revaz AI is introduced to clinical practice.

## Conclusions

This retrospective health economic simulation modeling study showed that AI-enabled clinical decision support can optimize many coronary revascularization decisions, leading to substantial cost savings and improved patient outcomes, even when limited clinicians’ AI adoption was assumed. The findings from this study demonstrate that the economic value of utilizing AI-based decision support can be substantial. Randomized clinical trials are warranted to validate the real-world impacts of Revaz AI on clinician decision-making, costs, and patient outcomes.

## Data Sharing Statement

The patient data used in this study contains real patient information and cannot be shared without permission from the data custodians, Alberta Health Services and Alberta Health. The simulated data may be shared upon reasonable request.

## Author Contributions

TM designed and executed the health economic simulation, produced all results, and wrote the manuscript. AP conceived the study and provided feedback on the study design. EB prepared the patient data and Revaz AI predictions for the simulation model. BH, CJM, RW, and BT provided clinical input. CLFS and JL provided technical input related to AI and clinical decision support. JL provided feedback on the study design, wrote the manuscript, and oversaw the project. All authors critically revised the manuscript.

