## Supplement 1 for "Health economic simulation modeling of an AI-enabled clinical decision support system for coronary revascularization"

### eFigures & eTables

Mullie et al.

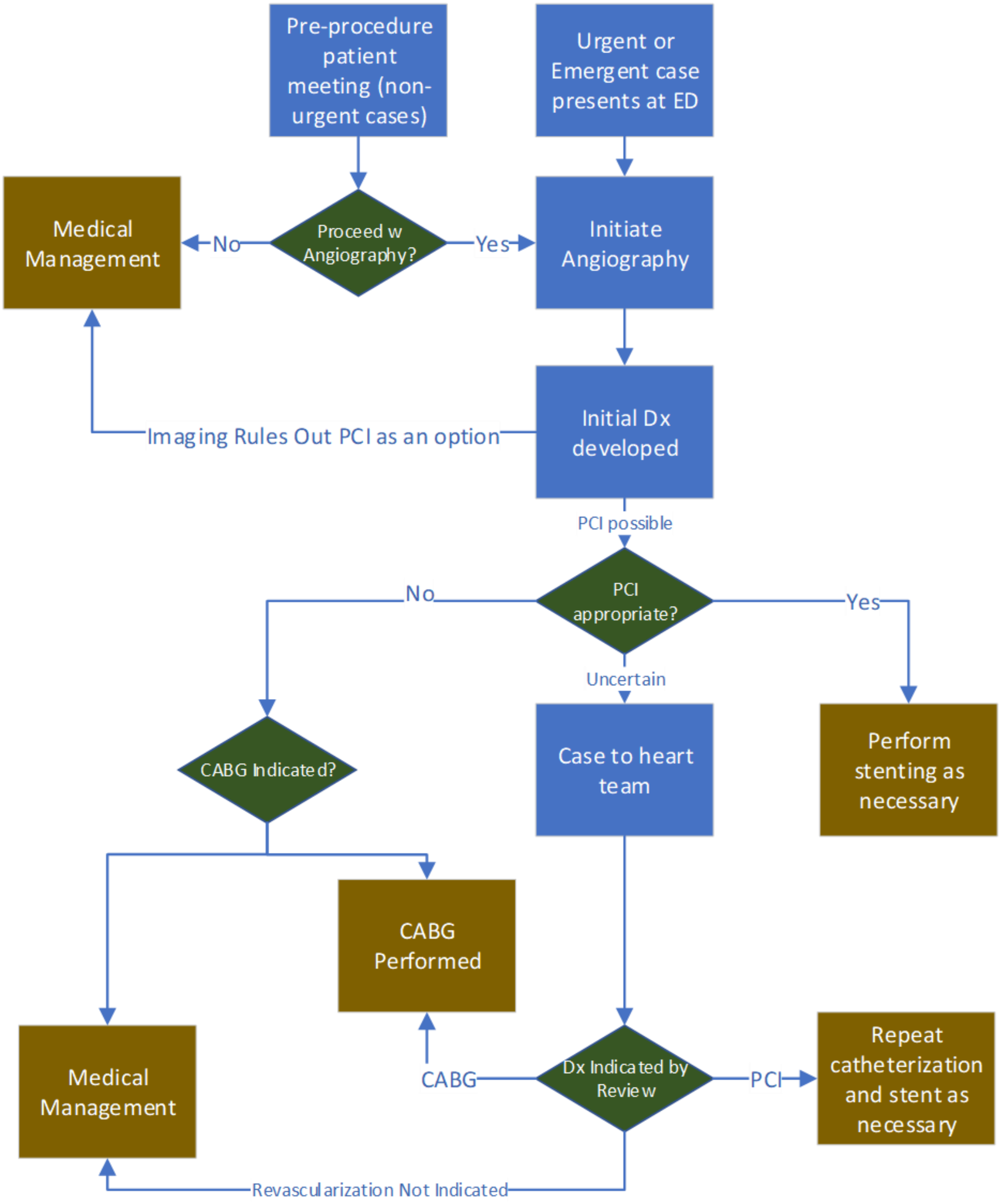


**eFigure 1**: Diagnosis and treatment flowchart for patients with coronary artery disease. In practice, this decision-making process can be complex. For example, non-urgent patients with stable angina might opt out of angiography entirely or opt to avoid CABG due to its invasiveness and significant recovery time. Patients may be found on the angiogram to be ineligible for some treatments, and in complex cases the interventional cardiologist may opt to end the diagnostic angiography without immediate PCI to consult with a heart team.

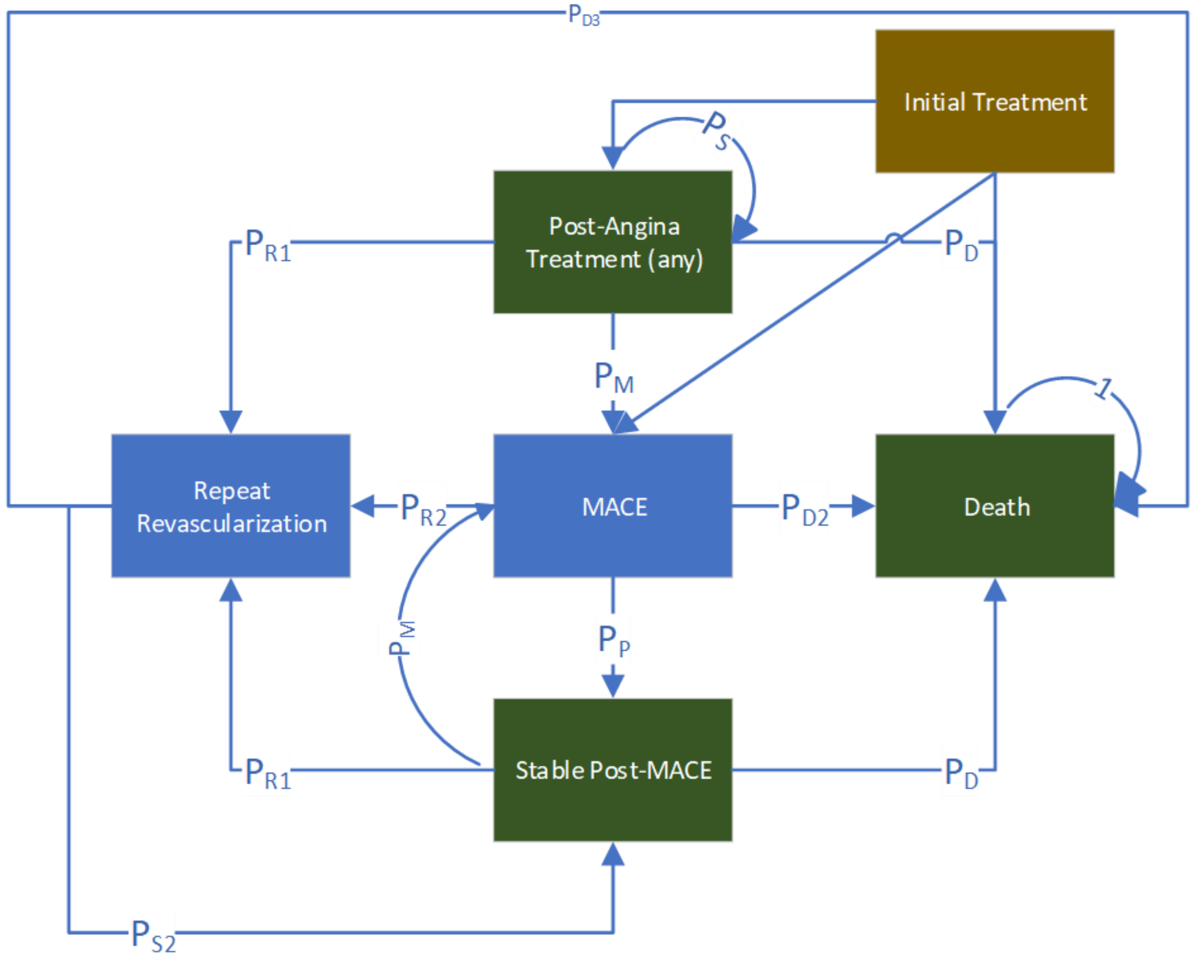


**eFigure 2**: Markov health economic simulation model structure.

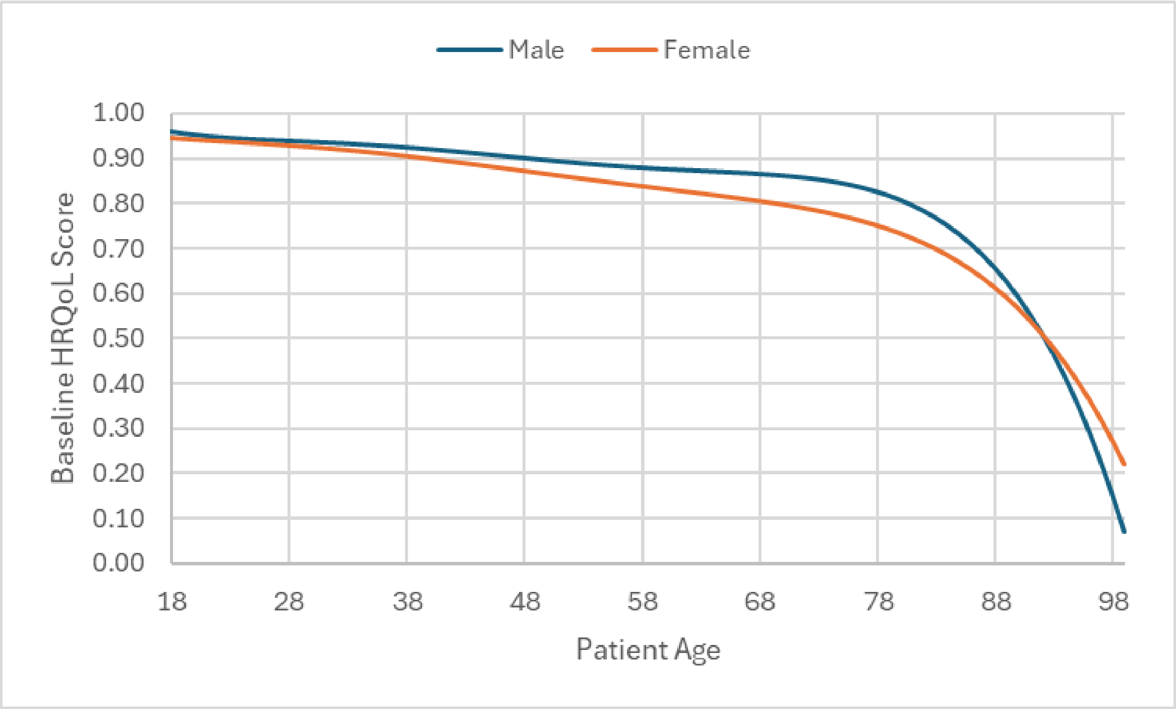


**eFigure 3**: Baseline health related quality of life as a function of patient age, stratified by patient sex.

**eTable 1**: Patient cohort characteristics used in the simulation model.

| Characteristics | Mean (standard deviation) or Percentage |
|---|---|
| Age (years) | 65.1 (11.5) |
| Male sex | 76% |
| Indication | |
| NSTEMI | 58% |
| Unstable Angina | 15% |
| Stable Angina | 27% |
| Diagnosis | |
| Left main (with or without other) | 13% |
| Multivessel (excluding left main) | 65% |
| Single vessel | 22% |

**eTable 2**: Event probabilities used in the simulation model, shown as annualized rates. Probability modifiers applied to individual Revaz AI probabilities are also shown. SV: single vessel, MV: multivessel (2 or more), LM: left main (with or without any other vessels occluded), MI: myocardial infarction, MACE: major adverse cardiovascular event, MT: medical therapy only, PCI: percutaneous coronary intervention, CABG: coronary artery bypass graft, SD: standard deviation.

| Variable | Value | Relative SD | Sources |
|---|---|---|---|
| Patient has a history of MI | 32.8% | 0.2 | SYNTAX trial[1] |
| Proportion of 3-year risk in cycle 1 | 30% | 0.2 | SYNTAX trial[1] |
| Probability of revascularization without a new MI<br>PCI (SV or MV)<br>MT (SV or MV)<br>CABG (SV or MV)<br>PCI (LM)<br>MT (LM)<br>CABG (LM) | <br><br>3.9%<br>5.3%<br>1.6%<br>11.4%<br>15.6%<br>5.4% | 0.2 | Systematic review[2]<br>COURAGE trial[3]<br>SYNTAX trial[4]<br>Systematic review[5] |
| Probability of revascularization following MI | 32% | 0.2 | FAME 2 trial[6] |
| Probability that revascularization during follow-up is PCI (vs. CABG) | 80% | 0.15 | SYNTAX trial[4] |
| Peri-operative or immediate post-operative death, PCI | 1.2% | 0.1 | Registry data[7] and SYNTAX trial[8] |
| Peri-operative or immediate post-operative death, CABG | 1.2% | 0.1 | Registry data[7] and SYNTAX trial[8] |
| Distribution of MACE in PCI:<br>MI<br>Stroke<br>Heart failure | <br>71.7%<br>16.7%<br>11.6% | 0.05 | ISCHEMIA trial[9] |
| Distribution of MACE in MT:<br>MI<br>Stroke<br>Heart failure | <br>73.5%<br>15.9%<br>10.6% | 0.05 | ISCHEMIA trial[9] |
| Distribution of MACE in CABG:<br>MI<br>Stroke<br>Heart Failure | <br>46.5%<br>42.9%<br>10.6% | 0.05 | SYNTAX trial data[8] adjusted to reflect heart failure prevalence in ISCHEMIA trial patients[10] |
| Lower bound mortality risk | 1.14% | N/A | Statistics Canada[11] |
| Lower bound MACE risk | 0.77% | N/A | Statistics Canada[12] |

**eTable 3**: Mean cost values used in the simulation model. All costs are in 2024 USD. SV: single vessel, MV: multivessel (2 or more), LM: left main (with or without any other vessels occluded), MI: myocardial infarction, MT: medical therapy only, PCI: percutaneous coronary intervention, CABG: coronary artery bypass graft.

| Cost | Value | Sources |
|---|---|---|
| Diagnostic catheterization for MT patients | $11,139 | FAME 2 trial,[6] adjusted relative to PCI cost in SYNTAX, FREEDOM and EXCEL trials[13] |
| Total episode cost for PCI, excluding stent cost, initial revascularization procedure | $22,130 | Average of costs from the SYNTAX, FREEDOM, and EXCEL trials[13] |
| Total episode cost for CABG as initial revascularization procedure | $39,428 | Average of costs from the SYNTAX, FREEDOM, and EXCEL trials[13] |
| Total episode cost for CABG as repeat revascularization | $49,032 | EXCEL trial[13] |
| Total episode cost for PCI as repeat revascularization | $20,863 | EXCEL trial[13] |
| Stent cost | $1,530 | European cost, adjusted to USD[14] |
| Number of stents placed:<br>SV<br>MV<br>LM | <br>1.0<br>4.1<br>2.4 | Average from the SYNTAX, FREEDOM, and EXCEL trials[13] |
| Ongoing management cost, MT:<br>Year 1<br>Year 2<br>Year 3+ | <br>$2,673<br>$2,673<br>$2,673 | PCI and CABG costs from the EXCEL trial[13]<br><br>MT cost assumed equal to PCI cost in year 3+ |
| Ongoing management cost, PCI:<br>Year 1<br>Year 2<br>Year 3+ | <br>$5,777<br>$2,678<br>$2,673 | |
| Ongoing management cost, CABG:<br>Year 1<br>Year 2<br>Year 3+ | <br>$12,069<br>$2,753<br>$2,806 | |
| Aspirin cost/day | $0.065 | Alberta health insurance reimbursement rates October 2023[15] |
| ACE inhibitor cost/day | $0.108 | |
| Beta blocker cost/day | $0.100 | |
| Calcium channel inhibitor cost/day | $0.107 | |
| P2Y12 inhibitor cost/day | $0.210 | |

| | | |
|---|---|---|
| Statin cost/day | $0.187 | |
| Acute MI treatment cost without revascularization | $12,688 | EXCEL trial[13] |
| Acute stroke treatment cost | $15,381 | Literature review[16] |
| Heart failure hospitalization cost | $15,449 | Literature review[16] |
| Post-stroke ongoing management cost | $2,107 | Literature review[16] |
| MI ongoing management cost | $2,399 | Swiss cohort study[17] |
| Heart failure ongoing management cost | $1,374 | Literature review[16] |

**eTable 4**: Health state utilities used in the simulation model. MI: myocardial infarction, PCI: percutaneous coronary intervention, CABG: coronary artery bypass graft.

| State | Utility Modifier |
|---|---|
| New onset stable angina | 0.86 |
| Ongoing stable angina | 0.87 |
| New onset unstable angina | 0.74 |
| Ongoing unstable angina | 0.74 |
| New MI | 0.82 |
| Stable with history of MI | 0.83 |
| New stroke | 0.81 |
| History of stroke | 0.79 |
| New heart failure | 0.73 |
| Ongoing heart failure | 0.73 |
| PCI treatment (first cycle) | 1.112 |
| Past PCI treatment | 1.099 |
| CABG treatment (first cycle) | 1.018 |
| Past CABG treatment | 1.099 |

**eTable 5**: Actual treatment compared to ‘sticky’ physician treatment decisions in Scenario 3. MT: medical therapy only, PCI: percutaneous coronary intervention, CABG: coronary artery bypass graft.

| | | **Actual Treatment** | | |
|---|---|---|---|---|
| | | **MT** | **PCI** | **CABG** |
| **Optimal Treatment** | **MT** | 2,581 | 188 | 28 |
| | **PCI** | 2,516 | 14,744 | 581 |
| | **CABG** | 782 | 1,865 | 3,320 |
| **Net Gain/Loss** | | -3,082 | +1,044 | +2,038 |

10. Gerber Y, Weston SA, Berardi C, et al. Contemporary Trends in Heart Failure With Reduced and Preserved Ejection Fraction After Myocardial Infarction: A Community

Study. *American Journal of Epidemiology*. 2013;178(8):1272-1280. doi:10.1093/aje/kwt109
