## Supplement 2 for "Health economic simulation modeling of an AI-enabled clinical decision support system for coronary revascularization"

### Simulation Methodology Details

Mullie et al.

eFigure 2 (Supplement 1) illustrates the simulation model structure. The history of MI prior to the current event was not available for the simulation and was randomly assigned in each model iteration. The first cycle of the model involved the patient’s initial treatment and follow-up. PCI and CABG patients had a risk of peri- or immediate post-operative death, while patients with MT were assumed to survive diagnostic catheterization. Patients could also experience a MACE or death from cardiac or non-cardiac causes in the first cycle.

Following initial treatment, surviving patients were assigned to a stable post-treatment state. From that state, in each subsequent cycle, they could require new or repeat revascularization without an MI (assumed to be due to new or recurrent stable angina), experience a MACE, remain stable, or die of non-cardiac causes. Overall MACE and death probabilities were individual and taken from the Revaz AI predictions for that patient and the selected treatment modality. Patients experiencing a MACE were randomly assigned to one of MI (which could also result in repeat revascularization), stroke, or hospitalization for a new onset of heart failure. Repeat revascularization probabilities were taken from the literature and varied depending on the patient’s initial treatment.

Patients assigned to MACE or to revascularization without MI also had a risk of death resulting from that event, in which case they were assigned directly to the death state at the conclusion of the current cycle. Individuals who survived a MACE were assigned back to the stable condition but with altered ongoing costs and quality of life (see next subsections).

Each patient's probability of death concurrent with another event was scaled based on their death risk in that cycle relative to the population average risk in that cycle:

$$P_{(\text{Death from MACE i,c})} = P_{(\text{Cardiac death})} \times P_{(\text{Death i,c})} / P_{(\text{Death c})}$$

$$P_{(\text{Peri-operative death i,c})} = P_{(\text{Peri-operative death})} \times P_{(\text{Death i,c})} / P_{(\text{Death c})}$$

where $P_{(\text{Death from MACE i,c})}$ is the probability of dying conditional on having suffered a MACE in that cycle, where i indicates that a risk is particular to an individual, and c indicates that it is particular to a model cycle. $P_{(\text{Cardiac death})}$ is the expected proportion of all deaths in the cohort based on literature reports of the proportion of PCI or CABG study patients whose proximate cause of death is MI, stroke, or heart failure. $P_{(\text{Peri-operative death})}$ is the population level probability of death either during a revascularization procedure or before discharge from hospital following such a procedure. $P_{(\text{Death i,c})}$ is all-cause death probability for that individual in a given model cycle, and $P_{(\text{Death c})}$ is the average death probability of all patients in that cycle. $P_{(\text{Death from MACE i,c})}$ and $P_{(\text{Peri-operative death i,c})}$ were capped such that their sum could not be greater than 95% of $P_{(\text{Death i,c})}$. The probability of death from other causes was calculated as:

$$P_{(\text{Death other i,c})} = P_{(\text{Death i,c})} - P_{(\text{Death from MACE i,c})} + P_{(\text{Peri-operative death i,c})}$$

so that the overall probability of all cause death in the cycle was preserved as initially forecasted by Revaz AI.

It was assumed that MACE, death, and non-MI-related revascularization rates were higher during the first model cycle, as observed in the Kaplan–Meier curves of SYNTAX[1] and other trials. A higher proportion of 3-year MACE and mortality risks was assigned to the first model cycle, covering initial treatment and short-term recovery. Remaining 3-year mortality and MACE probabilities were then divided evenly across the remaining 11 cycles in that period.

Probabilities from cycles 13 through 20 were calculated by subtracting the individuals' Revaz AI-derived 3-year probability from their 5-year probability and assigning the difference evenly across those 8 cycles. In some cases, Revaz AI's estimated 5-year probabilities of either mortality or MACE were similar to or marginally lower than the same individual's 3-year probabilities. To account for this, lower bound mortality probabilities were calculated based on the annual death risk for members of the Canadian general population aged 65-70, translated into a quarterly probability. Lower bound MACE risks were calculated based on the annual incidence of MI or stroke among Canadians aged 65-70, again translated into quarterly probabilities. Individuals faced the higher of either the lower bound risk or their individual predictions from Revaz AI in each cycle.

In cycles beyond 20, cycle 20 risks were repeated, adjusted based on the change in relative risk of mortality between the patient's starting and current age in the Canadian general population.[2] For example, the annual death probabilities for a 65-69 year old, 70-74 year old, and 75-79 year old in Canada are 1.14%, 1.77%, and 2.90%, respectively. An individual who entered the model at age 65 would have their cycle 20 death risk multiplied by 1.77/1.14 = 1.55 in cycle 21 when they would be 70, and 2.90/1.14 = 2.54 in cycle 41 when they would be 75. Death risk was set to 100% in individuals older than 100 years to account for the lack of quality risk data at extreme ages. This had a minimal impact on overall results, as the probability of surviving to 100 was very low for all patients.

The risk of revascularization without MI was based on initial treatment modality, occlusion type, and observed rates in the literature. Risks were assumed to be tripled in the first cycle. Individuals who required repeat revascularization without MI were returned to their previous state following treatment if they did not experience death in the short term.

In patients assigned to repeat revascularization, either with or without MI, treatment modality was randomly assigned to be one of either PCI or CABG. This assignment was random because updated Revaz AI predictions would depend on changes in patient health state between the initial and repeat treatments, which were not available.

Individual-level Revaz AI risks were consistent across model iterations, but all other parameters were randomly sampled from a normal distribution. Standard deviations were set relative to mean values; for example, a 30% probability with a relative standard deviation of 0.2 had a standard deviation of 6%. Time-related probabilities are reported as annual but were converted into quarterly by the model.
