## Supplement 3 for "Health economic simulation modeling of an AI-enabled clinical decision support system for coronary revascularization"

### CHEERS 2022 Checklist

| | Item | Guidance for Reporting | Reported in section |
|---|---|---|---|
| **TITLE** | | | |
| Title | 1 | Identify the study as an economic evaluation and specify the interventions being compared. | Title page |
| **ABSTRACT** | | | |
| Abstract | 2 | Provide a structured summary that highlights context, key methods, results and alternative analyses. | Abstract |
| **INTRODUCTION** | | | |
| Background and objectives | 3 | Give the context for the study, the study question and its practical relevance for decision making in policy or practice. | Introduction |
| **METHODS** | | | |
| Health economic analysis plan | 4 | Indicate whether a health economic analysis plan was developed and where available. | N/A |
| Study population | 5 | Describe characteristics of the study population (such as age range, demographics, socioeconomic, or clinical characteristics). | AI-enabled CDSS, Patient Cohort |
| Setting and location | 6 | Provide relevant contextual information that may influence findings. | AI-enabled CDSS, Patient Cohort |
| Comparators | 7 | Describe the interventions or strategies being compared and why chosen. | Health Economic Simulation Model |
| Perspective | 8 | State the perspective(s) adopted by the study and why chosen. | Health Economic Simulation Model |
| Time horizon | 9 | State the time horizon for the study and why appropriate. | Health Economic Simulation Model |
| Discount rate | 10 | Report the discount rate(s) and reason chosen. | Supplement 2 |
| Selection of outcomes | 11 | Describe what outcomes were used as the measure(s) of benefit(s) and harm(s). | Supplement 2 |
| Measurement of outcomes | 12 | Describe how outcomes used to capture benefit(s) and harm(s) were measured. | Supplement 2 |
| Valuation of outcomes | 13 | Describe the population and methods used to measure and value outcomes. | Utilities |
| Measurement and valuation of resources and costs | 14 | Describe how costs were valued. | Costs |
| Currency, price date, and conversion | 15 | Report the dates of the estimated resource quantities and unit costs, plus the currency and year of conversion. | Supplement 2 |
| Rationale and description of model | 16 | If modelling is used, describe in detail and why used. Report if the model is publicly available and where it can be accessed. | Supplement 2 |
| Analytics and assumptions | 17 | Describe any methods for analysing or statistically transforming data, any extrapolation methods, and approaches for validating any model used. | Supplement 2 |
| Characterizing heterogeneity | 18 | Describe any methods used for estimating how the results of the study vary for sub-groups. | Supplement 2 |
| Characterizing distributional effects | 19 | Describe how impacts are distributed across different individuals or adjustments made to reflect priority populations. | Supplement 2 |
| Characterizing uncertainty | 20 | Describe methods to characterize any sources of uncertainty in the analysis. | Supplement 2 |
| Approach to engagement with patients and others affected by the study | 21 | Describe any approaches to engage patients or service recipients, the general public, communities, or stakeholders (e.g., clinicians or payers) in the design of the study. | N/A |
| **RESULTS** | | | |
| Study parameters | 22 | Report all analytic inputs (e.g., values, ranges, references) including uncertainty or distributional assumptions. | Supplement 2 |
| Summary of main results | 23 | Report the mean values for the main categories of costs and outcomes of interest and summarise them in the most appropriate overall measure. | Results |
| Effect of uncertainty | 24 | Describe how uncertainty about analytic judgments, inputs, or projections affect findings. Report the effect of choice of discount rate and time horizon, if applicable. | Results |
| Effect of engagement with patients and others affected by the study | 25 | Report on any difference patient/service recipient, general public, community, or stakeholder involvement made to the approach or findings of the study | N/A |
| **DISCUSSION** | | | |
| Study findings, limitations, generalizability, and current knowledge | 26 | Report key findings, limitations, ethical or equity considerations not captured, and how these could impact patients, policy, or practice. | Discussion |
| **OTHER RELEVANT INFORMATION** | | | |
| Source of funding | 27 | Describe how the study was funded and any role of the funder in the identification, design, conduct, and reporting of the analysis | Funding |
| Conflicts of interest | 28 | Report authors conflicts of interest according to journal or International Committee of Medical Journal Editors requirements. | Conflicts of Interest |

Husereau D, Drummond M, Augustovski F, de Bekker-Grob E, Briggs AH, Carswell C, Caulley L, Chaiyakunapruk N, Greenberg D, Loder E, Mauskopf J, Mullins CD, Petrou S, Pwu RF, Staniszewska S; CHEERS 2022 ISPOR Good Research Practices Task Force. Consolidated Health Economic Evaluation Reporting Standards 2022 (CHEERS 2022) Statement: Updated Reporting Guidance for Health Economic Evaluations. BMJ. 2022;376:e067975.
The checklist is Open Access distributed in accordance with the terms of the Creative Commons Attribution (CC BY 4.0) license, which permits others to distribute, remix, adapt and build upon this work, for commercial use, provided the original work is properly cited. See: http://creativecommons.org/licenses/by/4.0/.
